# Women’s perceptions of food safety risks and vendor practices in Northern Tanzania: a mixed-methods analysis

**DOI:** 10.64898/2025.12.17.25342242

**Authors:** Nishmeet Singh, Quinn Marshall, Lilia Bliznashka, Rachel Zakayo, Lydia Mukasa, Jose Luis Torres Chavez, Evangelista Malindisa, Kidola Jeremiah, Joyce Kinabo, Alexandra L. Bellows, Deanna K. Olney, Mercy Mwambi, Marisa Wilson, Lindsay M. Jaacks, Neha Kumar

## Abstract

We know little about how women in Africa perceive and manage food safety risks for fruits and vegetables, and how these perceptions and practices influence their food choices. To investigate this, we employed a sequential mixed-methods analysis using data from 33 villages across the Arusha and Kilimanjaro regions of Northern Tanzania. Quantitative household surveys were conducted with 2,577 women to document their perceptions of food safety risks associated with fresh fruits and vegetables. Additionally, quantitative surveys assessing food safety facilities and practices were conducted in 14 markets and 108 retail outlets. These data informed the creation of food safety scores for women’s risk perceptions and village-level vendor characteristics. We used the scores to analyse their relationship with household-level consumption and expenditure on fruits and vegetables using mixed-effects regression models. Subsequently, ethnographic research involved in-depth exploration with women from three communities to investigate the factors shaping food safety risk perceptions and their connections to household practices and food choices. Our quantitative findings indicated that women’s perceptions of food safety were associated with household-level consumption and expenditure, while vendor safety characteristics were not. The qualitative insights revealed that social relations, community interactions, and trust were pivotal for women in managing food safety risks. Women used these to assess and address food safety risks. Women were also assisted in minimising risk through their roles as producers, sellers, and household managers. Our analysis highlights the importance of understanding consumer perceptions and their socio-cultural contexts in designing interventions that enhance food safety and promote healthier diets.

## 1. Introduction

Nutrient-dense fresh fruits and vegetables are essential for healthy, diverse diets; however, they often pose a high risk of contamination, which can lead to foodborne diseases (FBDs) or illnesses ranging from diarrhoea to cancer (Lambertini et al., 2024; World Health Organization, 2022). Foodborne hazards also reduce the nutritional value of fruits and vegetables, increasing the risk of malnutrition (Khan et al., 2015). The burden of FBDs is elevated in several African and Asian countries because fresh fruits and vegetables for domestic consumption are primarily produced, handled, transported, and sold by people with limited economic and operational capacity, such as limited capital to invest in safer equipment, hygiene and sanitary facilities, and protective utensils (Grace, 2023; Wallace et al., 2022). Further, domestic agrifood systems in Africa are weak in safety-related infrastructure and regulatory support, such as clean water, waste disposal facilities, credit facilities, public food safety advisors and training, and testing labs (Ayalew et al., 2023; Grace, 2023; Jaffee et al., 2020). Despite the challenges, there is also evidence of vendors’ positive attitudes and motivation to sell safe products in these contexts (Garsow et al., 2025; Lee et al., 2022; Roesel & Grace, 2014; Wenndt et al., 2025).

The World Health Organisation (WHO) defines food safety as "…limiting the presence of those hazards, whether chronic or acute, that may make food injurious to the health of the consumer" (WHO, Regional Office for South-East Asia, 2015, p. 5). Food safety influences food choices at multiple socio-ecological levels, from individual (wealth, experience of FBDs, food familiarity, perceptions, prior knowledge, etc.), social (norms, vendor reputation, culture, etc.), physical (sanitation and hygiene in and around outlets, food presentation, home-gardens, etc.), to macro environments (regulations, technical capacity, etc.) (Liguori et al., 2022; Osei-Kwasi et al., 2020). At the individual or consumer level, food safety and food choices are linked through perceptions and context (Lambertini et al., 2024; Nordhagen, Lambertini, et al., 2022). Food safety risk perceptions are an individual’s understanding of the health or disease risks associated with consuming unsafe food. Subjective and psychosocial factors, such as risk perceptions, are associated with people’s willingness to buy food and with their safe food-handling behaviour, such as practising personal hygiene or avoiding risky foods (Machado Nardi et al., 2020; Young et al., 2017).

Data from 2010 indicate that countries in the East African region had a per capita FBD burden of 1,200 disability-adjusted life years (DALYs) per 100,000 population compared to a global average of 477 DALYs (Havelaar et al., 2015). In the region, food safety perceptions are influenced by the presence of microbial and chemical hazards identified in farm-level risk assessments and in food environments (Kapeleka et al., 2020; Mutua *et al*., 2021; Roesel & Grace, 2014). It has been suggested that the intensive use of agrochemicals, such as industrial fertilisers and pesticides, has increased concentrations of toxic compounds above the maximum permissible limits on East African agricultural farms (Mng’ong’o et al., 2021). High microbial contamination and elevated levels of heavy metal and pesticide residues in multiple fresh vegetables have also been documented (Fuhrimann et al., 2021; Mutua et al., 2021). Finally, multiple systematic reviews, including those in East Africa, have shown that pesticide exposure, heavy metal pollution, and environmental contaminants are associated with pregnancy complications, adverse birth outcomes, poor child growth and development, and chronic diseases among adults (Chowdhury et al., 2018; Hassen et al., 2024; Lin et al., 2023; Udom et al., 2025).

In Tanzania, rural households are increasingly producing foods, including fresh fruits and vegetables, for sale, while also purchasing them. Evidence suggests that excessive use of agrochemicals at the farm level raises consumer concerns about their negative effects on human health (Sheahan et al., 2017). Beyond the farmgate, vendors’ practices relating to food safety are likely to alter consumer perceptions, practices, knowledge, and behaviours, resulting in changes in the types of food purchased, food sources, time spent and behaviours in the market, and at-home practices (Constantinides et al., 2020; Karanja et al., 2022; Stadlmayr et al., 2023; Wallace et al., 2022). Context-specific studies on ’how and why’ food safety is critical to fruit and vegetable choices among those living in the villages remain limited.

Women are primarily responsible for fruit and vegetable farming, selling, purchasing, and household consumption in Tanzanian villages (Akram-Lodhi, 2024; Kavishe, 1991; Keding et al., 2011; van der Maden et al., 2021). A notable research gap is the underrepresentation of women in empirical and mixed-methods research on fresh food systems and food safety (de Kanter *et al*., 2024; Grace, 2023; Lee et al., 2022; Stadlmayr et al., 2023). In particular, evidence is sparse regarding how their food safety experiences, including perceptions, beliefs, practices, and vendor interactions, influence decision-making for fruits and vegetables. Food safety of fruits and vegetables has implications for women’s health, diets, livelihoods, social participation, and household labour (Grace, 2023).

This analysis examined food safety for fruits and vegetables in northern Tanzania using a mixed-methods approach. Our quantitative research described the vendors’ food safety characteristics and women’s perceptions of food safety risk and then tested associations with household consumption and expenditure of fruits and vegetables. We hypothesised that women’s perceived risks to food safety would negatively influence outcomes, while vendors’ adherence to good food safety practices would positively influence them. Our qualitative research investigated how women perceive and practice food safety in their families, communities, and food environments. We used ethnographic research methods to present a contextual understanding of the determinants of women’s perceptions of food safety and explored potential links to household-level food safety practices and food-choice behaviours.

## 2. Methods

### 2.1 Setting

This analysis is based on data from the Arusha and Kilimanjaro regions of Northern Tanzania, two of the largest producers of fruits and vegetables in the country (Hadija et al., 2023; Kimaro, 2019). Significant levels of pesticide residues and microbial contamination were found in farms and markets supplying fresh fruits and vegetables in the two regions (Kapeleka et al., 2020; National Bureau of Statistics (NBS), 2025).

### 2.2 Sequential mixed methods design

We drew on baseline data from a cluster-randomised controlled trial being conducted in 33 villages across five districts in the two regions (Bliznashka et al., 2023). The trial aims to evaluate the impact of an integrated package of supply-side, demand-side, and food environment interventions on household vegetable production and women’s fruit and vegetable consumption. Women were eligible to participate if they were aged 15–49 years, had at least one biological child aged 10–14 years living in the same household, and intended to remain in the study area for the trial’s duration.

We followed a mixed-methods approach in this paper. First, we used quantitative data from the trial, which were collected at two levels between October 11, 2023 and January 15, 2024: household-level data from 2,604 women and food environment–level data from 108 single vendor retail outlets and 14 open-air markets at the ward level (an administrative division in Tanzania between region and district) selling fruits and vegetables. Then, we drew on qualitative research conducted between February and March 2024 in three villages in the Arusha region, purposively selected out of the 33 villages in the trial.

### 2.3 Quantitative component

#### Data collection: food environment and household

We employed a purposive sampling approach to select retail outlets and markets, based on key informant input and a food environment census. Assessment of retail outlets and markets covered food safety facilities and vendor food-handling practices using a contextually adapted list for the safety of fruits and vegetables (Wallace et al., 2022). The number of retail outlets in a village ranged from 0 to 6. Ward-level markets were common across multiple villages.

The food safety facilities data were collected through enumerator observations of five facilities in and around retail outlets and markets (Table 1). The vendor food-handling data used a combination of observations and vendor self-reporting for 10 practices. In markets, one vendor was randomly sampled separately for fruits, leafy vegetables, and non-leafy vegetables. Whereas, among retail outlets, the food-handling data was collected for combined fruits and vegetables.

**Table 1:** Details about the mixed-methods data collection

| Research component | Level | Gathered information | Data collection period |
| --- | --- | --- | --- |
| **Quantitative** | Market survey at the food environment level | Food safety facilities - observed: waste management, water access, food and handwashing facilities, toilet availability, and sewage systems | October 2023 |
|  |  | Food safety handling practices- observed: garbage around the outlet display, dirty water around the display, items displayed on the ground, dust or dirt on items, dust or dirt around items, items exposed to rain, items exposed to sun; self-reported: items moistened by water in an open vessel or jar, items moistened by water in a closed spray bottle, and the sale of pre-cut or peeled items<br>Collected separately for fruits, leafy vegetables, and non-leafy vegetables | November 2023 |
|  | Retail outlet survey at the food environment level | Food safety facilities - observed: waste management, water access, food and handwashing facilities, toilet availability, and sewage systems | October 2023 |
|  |  | Food safety handling practices - observed: garbage around the outlet display, dirty water around the display, items displayed on the ground, dust or dirt on items, dust or dirt around items, items exposed to rain, items exposed to sun; self-reported: items moistened by water in an open vessel or jar, items moistened by water in a closed spray bottle, and the sale of pre-cut or peeled items<br>Collected for combined fruits and vegetables | October 2023, November 2023, January 2024 |
|  | Women's survey at the household level | Food safety risk perception module- self-reported:<br>Perceived control: six statements on a 5-point Likert scale (Completely disagree to Completely agree)<br>Perceived concerns: five statements on a 6-point Likert scale (No concern to Concern, and do not know)<br>Collected separately for fruits and vegetables | October 2023 to January 2024 |
| **Qualitative** | Community and village level | • The three village communities were selected based on the number of fruit and vegetable sources available and geographically accessible to them, as well as their proximity to the nearest town<br>• In each village, we conducted observations in the community and food environments, including of the retail outlets and the weekly ward-level market accessible to them.<br>• Informal conversations were conducted with village development officers, agricultural extension officers, community leaders, and women's development groups. | January 2024 to March 2024 |
|  | Women level | • In each village, women were selected for the in-depth interviews based on household wealth, food sources accessed by them, and their primary occupation in the last 12 months.<br>• Understanding food safety was part of a broader qualitative investigation into women's perspective on the drivers of food choices in the communities. The interview guide covered food safety perceptions and practices of women when buying fruits and vegetables, related learning mechanisms, perceived consequences, and strategies to minimise risks. | January 2024 to March 2024 |

Women answered a food safety risk perception module consisting of six statements on a 5-point Likert scale for perceived risk control, and five statements on a 6-point Likert scale for perceived concerns (Table 1) (Wertheim-Heck & Raneri, 2020). Perceived risk control and concerns are two known antecedents of food safety risk perceptions (Machado Nardi et al., 2020).

The household head answered a separate module on household food consumption and expenditures, which recorded quantities consumed, purchased, produced at home, and from other sources, such as gifts, for the seven days preceding the survey, covering 146 items typically consumed in Tanzania. This analysis utilised the value of the total food consumption (purchased, home production, gifts, and other sources) and purchased food expenditure. We also collected demographic and socio-economic data for all households included in the sample. Separate teams collected data at the household and food environment levels. Enumerators from both teams were trained on the questionnaires for two weeks prior to the start of data collection

#### Measures

Our two key exposures of interest were the vendor food safety characteristics and women’s food safety risk perceptions. For the former, we created a village-level vendor food safety score (range: 0-24) with a higher score indicating better observed food safety for fruits and vegetables at the village level. Washing items removes pathogens; it depends on water quality (Augustsson et al., 2023). Although we collected information on whether vendors moisten fruit and vegetables or pre-cut items, we did not use these in the score calculation because of insufficient information on water quality and vendor handling practices for pre-cut items in the study regions. We initially calculated a total score for food safety facilities (range: 0-5) and practices (range: 0-7), separately for retail outlets and markets. Because there were multiple retail outlets in each village, we calculated the village median for the facilities and practices scores. Since markets are at the ward level, the scores were mapped one-to-one to the villages. We then summed the scores from the two sources to obtain a combined vendor food safety score.

We created an individual-level variable for the food safety risk perception score (range 12-110), with higher scores indicating lower perceived risk among women, implying better perceived food safety. The score was calculated as the sum of responses to 11 statements (Lai et al., 2021; Spagnoli et al., 2023). We reverse-coded the concern scale before summing it because it is positively associated with risk perceptions, whereas perceived control is negatively associated with risk perceptions (Machado Nardi et al., 2020). Then, for regression analysis, we used the tertiles of women’s risk perception scores to create a categorical variable indicating low, medium, or high risk perception levels. Similarly, we created levels for the vendor’s food safety score: low, moderate, or high.

We estimated household monthly per-capita consumption and expenditures in Tanzanian Shillings (TZS) for combined fruits and vegetables by dividing the total value by household size. This estimate served as a proxy for individual-level availability (Choudhury et al., 2020). We replaced eight outliers in consumption and expenditures with reported values >3 SD above or below the mean, using the median values of their respective villages.

#### Data analysis

We generated descriptive statistics for vendor safety characteristics in markets and retail outlets, as well as for women’s food safety risk perception statements, using counts and frequencies. In addition, we reported summary statistics on combined fruit and vegetable consumption and purchases in TZS per person per month.

We first tested the correlation between vendor food safety scores and women’s food safety risk perception scores using Kendall’s rank correlation tau (τ), which is a non-parametric measure robust to non-normality and tied values in the data (Okoye & Hosseini, 2024). The complete case analysis was used to handle missing data (<1% across all variables), yielding a sample of 2,577 households for regression analysis.

We used linear mixed-effects models to investigate whether women’s perceived food safety risk and vendor food safety were associated with fruit and vegetable consumption and expenditures. We used a village-level random-intercept model to account for clustering in the data of the form:

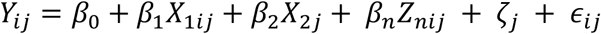

where 𝑌_𝑖𝑗_ is the dependent variable for household or individual i in village j village, 𝑋_1𝑖𝑗_ is the food safety risk perception level for individual i in village j, 𝑋_2𝑗_ is the vendor food safety score in village *j,* 𝑍_𝑛𝑖𝑗_ are the covariates for household or women i in village *j,* 𝜁_𝑗_ ∼ N(0, ψ) is a village-specific random intercept and 𝜖_𝑖𝑗_ is the individual-level error term. We report regression coefficients, along with their 95% confidence interval.

We adjusted the regression models for a set of potential confounders identified using a Directed Acyclic Graph (DAG) based on published studies (Ha et al., 2020; Machado Nardi et al., 2020; Rashid et al., 2024) and further modified after discussion among co-authors (Supplemental Figure 1). The final confounders were household wealth (calculated using a principal component analysis of 12 assets and seven housing characteristics), household dependency ratio, household production of any fruit or vegetable in the last 12 months, and women’s age, education, and primary occupation.

The quantitative analyses were conducted using R version 4.5.0 (R Core Team, 2025). Data cleaning was done using STATA version 18 (StataCorp, 2023).

### 2.4 Qualitative component

#### Data collection

We used a mixed-method approach to add contextual depth to the quantitative findings of food safety and its relationship with food consumption measures. For the qualitative research, we implemented a purposive maximum variation sampling approach (Palinkas et al., 2015) to select villages and women based on quantitative data from the FRESH survey, thereby capturing differences and similarities in women’s social and cultural lived experiences of food safety within and across villages (Table 1).

We employed ethnographic research methods to generate data through detailed and engaged fieldwork in the village communities (Coffey, 2018). This included a combination of observations, informal conversation, in-depth interviews, listening, and reflecting. In-depth interviews were conducted with 20 women across the three villages. The participants were interviewed about their perceptions and practices of food safety in their families, communities, and food environments.

The authors initially conducted a desk review of available published and grey literature to develop an English interview guide, which was then translated into Kiswahili by the research assistants (RZ and LM). The research assistants were from Tanzania with prior experience conducting field surveys with women and on food-related topics. The lead author (NS) and the research assistants spent between one and two weeks discussing the research design and interview guides in a classroom setting, followed by observations in each community before the interviews. We also met respondents at their homes to schedule interviews and observe their surroundings, including gardens and farms. The pre-visits and observations helped us revise the interview guide, build rapport with the participants, and become accustomed to the context (Guest et al., 2013).

The in-depth interviews were conducted in Kiswahili at the participants’ homes by research assistants in the presence of the lead author. The interviews were also audio-recorded with the women’s consent, and the recordings were later transcribed and translated into English (RZ and LM). At the end of each interview, we asked participants about their willingness to participate in a shorter follow-up conversation scheduled for 3-

4 weeks after the initial interview. This step allowed us to gather early insights from the interviews to confirm, enrich, or transform the point of inquiry. We conducted follow-up conversations with 10 women to share preliminary insights, avoid misinterpretation, and allow the women to present any disagreements (Buscatto, 2018).

#### Data analysis

We used the reflexive thematic analysis (RTA) for the data analysis by incorporating our observations and experiences in the communities during the discovery of themes in the interview data, while simultaneously reflecting on our process, practices, and perspectives throughout data collection, analysis, and interpretation (Braun, 2022; Byrne, 2022). Our interpretative framework was constructivist with an experiential orientation. As such, we realised that food safety reflected women’s social or lived experiences and then described it through themes as experienced by the participants. Based on the analytic process for RTA, we familiarised ourselves with the data by reading all transcripts independently and then discussing each transcript as a group. We also took brief notes for each interview and used them during the follow-up interviews. The interview data analysis began with re-reading the transcripts independently, followed by hand-written descriptive coding by researchers (NS, RZ, LM), and then axial thematic coding along our initial line of inquiries, such as learning mechanisms or participants’ strategies to minimise risks (Cope & Kurtz, 2024). Here, we also focused on data sequences by including questions or relevant prior responses (Silverman, 2019). We then entered the codes into the analysis software, and the researchers (NS and RZ) worked independently to generate initial themes. The codes were later reviewed, revised and developed (by NS) into the core themes aligned with the research objectives. The qualitative software Dedoose, version 10.0.25, was used to code and develop themes.

## 3. Findings

### 3.1 Quantitative

#### Vendor food safety characteristics

The observed markers of food safety facilities or infrastructure were similar at the market and retail outlet levels (Figure 1 and Supplemental T1). A toilet facility was observed in most cases (market: 93%, retail outlet: 98%), followed by a dedicated waste collection area (market: 93%, retail outlet: 82%). The least common facility was a hand-washing or food-handling station (market: 0%; retail outlet: 30%). Both markets and retail outlets had a median safety facility score of 3 out of 5 (Q1 - Q3, 2 - 4).

**Figure 1:**
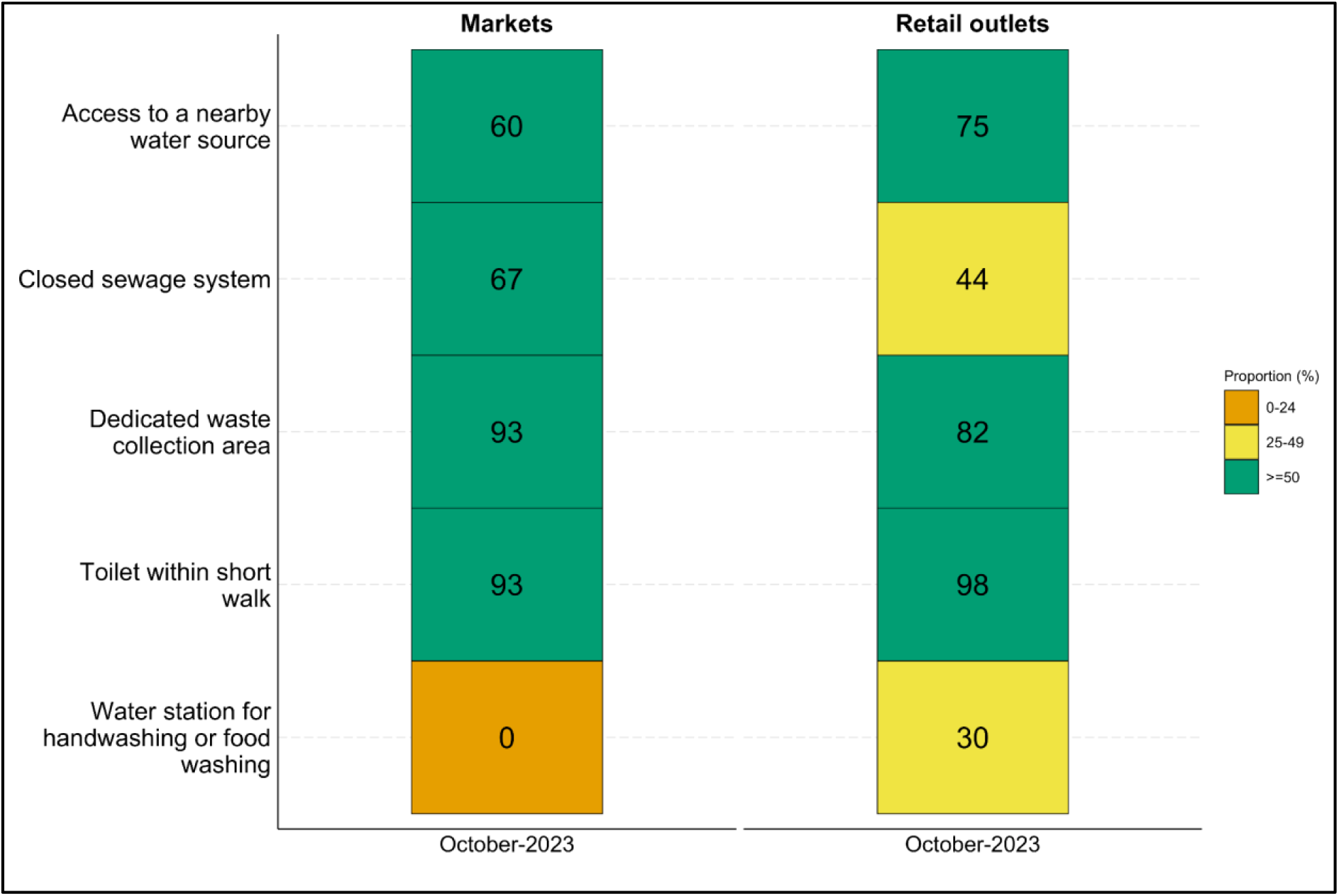
Proportion of vendors with food safety facilities at the market (n=15) and retail outlet level (n=108) in October 2023

In-terms of fruit and vegetable handling practices at the markets (Figure 2 and Supplemental T2), a high proportion of garbage around the display (fruit: 54%, leafy vegetables: 62%, non-leafy vegetables: 43%) and items displayed on the ground (fruit: 46%, leafy vegetables: 39%, non-leafy vegetables: 57%) was observed. In less than one-third of markets, we observed that fruit and vegetables were exposed to direct rain (fruit: 31%; leafy vegetables: 23%; non-leafy vegetables: 29%) or sunlight (fruit: 31%; leafy vegetables: 23%; non-leafy vegetables: 36%). The median market safety practice score for combined fruit and vegetables was 4.6 out of 7 (Q1 - Q3, 3.3 - 6).

**Figure 2:**
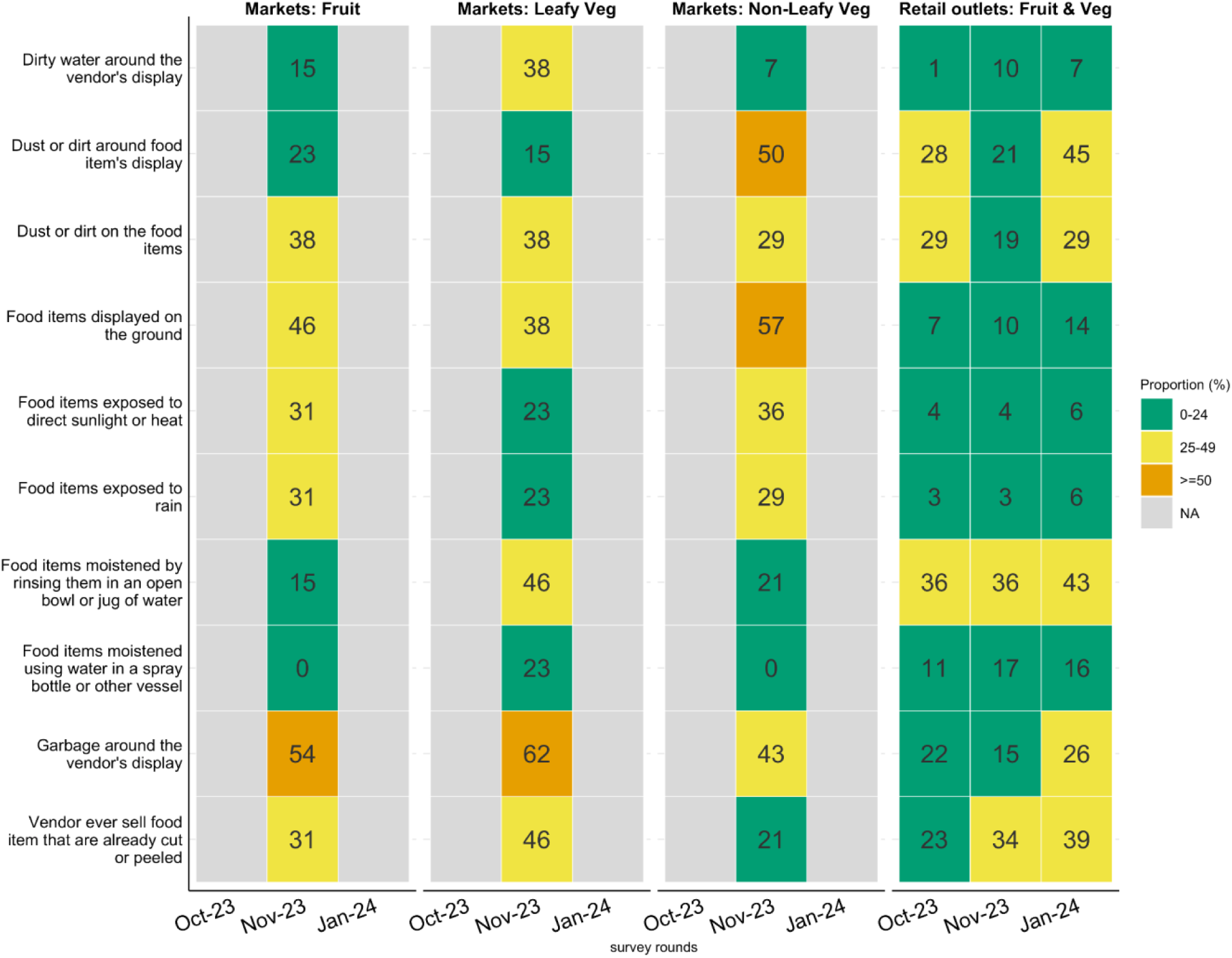
**Proportion of vendors with fruit and vegetable safety practices at the markets (n=14) and retail outlets (n=107) across survey rounds**

At the retail outlet level, on average across the three survey rounds, a low proportion of vendors displayed fruit and vegetables exposed to direct rain (4%) or sunlight (5%) (Figure 2 and Supplemental T2). A relatively higher proportion of vendors had dust or dirt on (31%) and around (26%) items, as well as garbage around the outlet (21%). The median safety practice score for retail outlets across the three survey rounds was 6 out of 7 (Q1 - Q3, 5 - 7).

#### Demographic characteristics and women’s food safety risk perceptions

The mean age of women was 38 years (SD 6.2). On average, 73% had attended or completed primary education, 86% were married, 59% were engaged in farming or livestock rearing, and 25% were involved in non-food-related activities (non-farm labour, non-food business or salaried work) (Supplemental T3). Women lived in households with an average of 6 (1.7) members, and 37% had produced any fruit or vegetable in their own farms or home-gardens in the 12 months preceding the survey.

Most women agreed that they only buy fruits and vegetables from vendors that use hygienic and protective measures (fruit: 91% agreed or completely agreed, vegetables: 92%) and that they only select items that are uncut or have peels (fruit: 92% agreed or completely agreed, vegetables: 91%, Figure 3 and Supplemental T4). Very few women agreed that they only select fruit or vegetables with certification or labels (fruit: 18% agreed or completely agreed, vegetables: 13%). Women primarily reported concerns about pesticide use (fruit: 56%, totally concerned or concerned, vegetables: 78%) and unhygienic vendor handling practices (fruit: 69%, totally concerned or concerned, vegetables: 75%, Figure 4 and Supplemental T4). Overall, women’s median food safety risk perception score was 58 out of 110 (Q1- Q3, 52 - 65).

**Figure 3:**
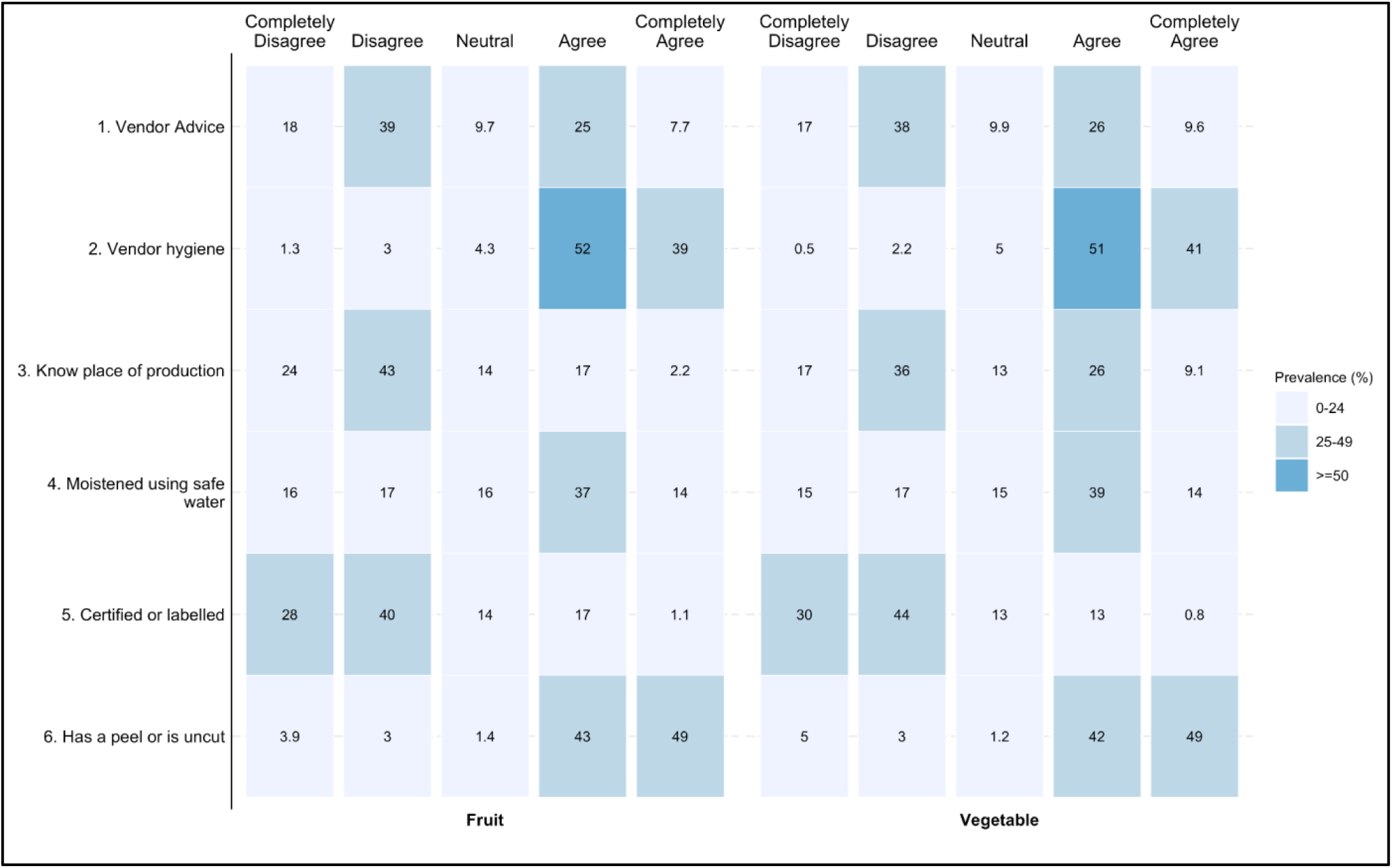
**Women’s description of fruit and vegetable safety risks based on statements related to perceived risk perceptions (n=2,593)**

**Figure 4:**
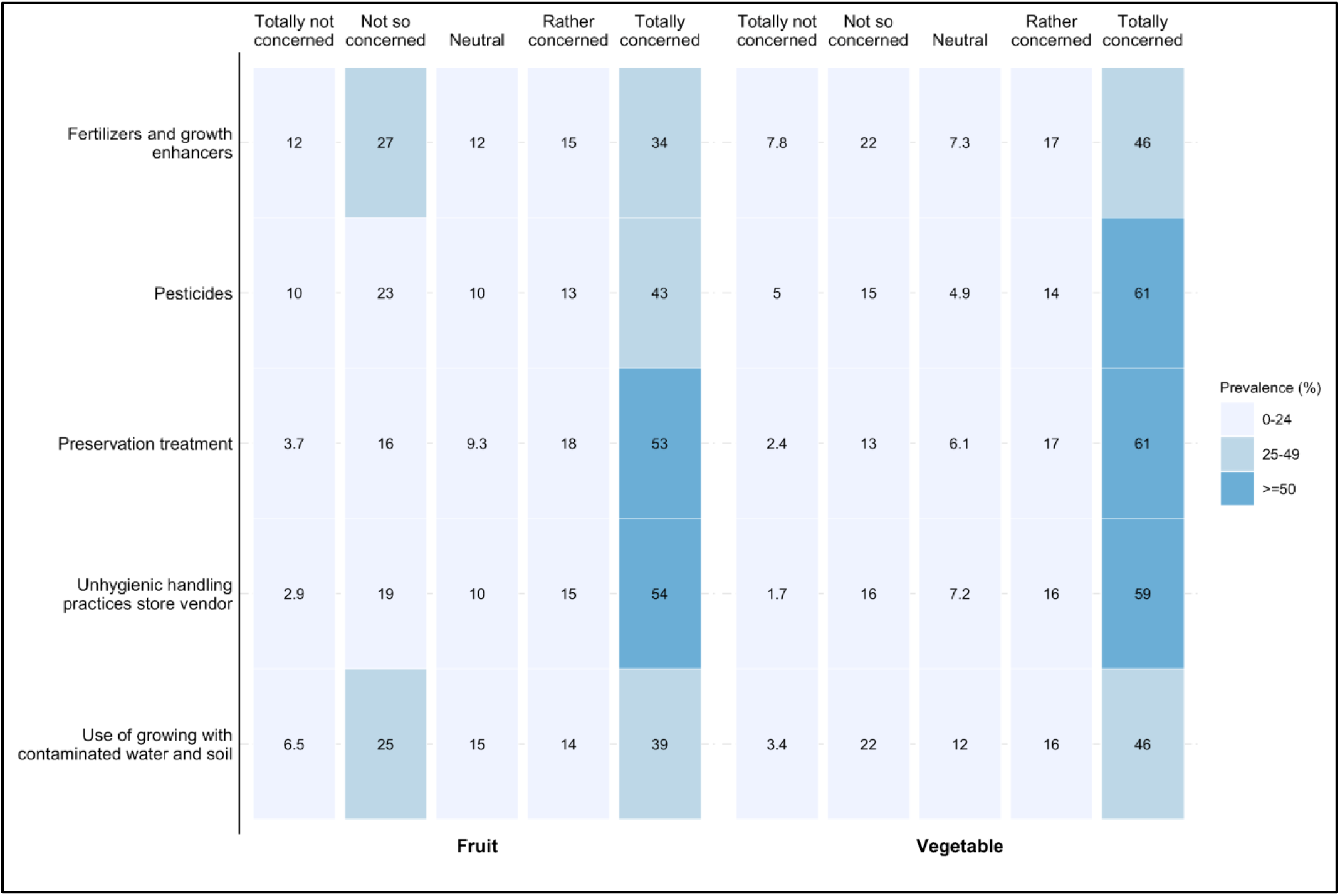
**Women’s description of fruit and vegetable safety risks based on statements related to perceived concerns (n=2,593)**

#### Correlation and Regression Results

The median monthly per capita consumption of fruit and vegetables was TZS 9,283 (Q1 - Q3: 5,138 - 14,877), while expenditures were Tanzanian Shillings 5,866 (3,205 - 9,622) (Supplemental T5). The correlation analysis showed no significant association between women’s food safety risk perceptions and vendor food safety characteristics (τ = -0.012, p = .358) (Supplemental T6). The results from the adjusted mixed-effects models indicated that fruit and vegetable consumption (person/month) for women with medium or low food safety risk perceptions, respectively, were greater by TZS 1,000 (95% CI: 194, 1,805) and TZS 832 (95% CI: 26, 1,637), compared to women with high food safety risk perceptions (Table 2, Supplemental T7 and T8). Similarly, fruit and vegetable expenditure (person/month) for women with medium or low food safety risk perceptions, respectively, were also greater by TZS 960 (95% CI: 455, 1,465) and TZS 772 (95% CI: 266, 1,279) compared to those who perceived high food safety risks. Vendor food safety was not associated with either consumption or expenditures. Other significant factors for fruit and vegetable consumption were household wealth, total per capita monthly food consumption, and the household’s production of fruits and vegetables in the last 12 months.

**Table 2:** Association between women’s food safety risk perceptions, vendor food safety characteristics and fruits and vegetables consumption and expenditure (TZS) (n=2,577)

|  | **Fruits and vegetables consumption**<br>**(1)** |  | **Fruits and vegetables expenditure**<br>**(2)** |  |
| --- | --- | --- | --- | --- |
| **Independent variable** | **Beta** | **95% CI** | **Beta** | **95% CI** |
| Women's risk perceptions |  |  |  |  |
| High | Ref | - | Ref | - |
| Medium | 1,000 | 194, 1,805 | 960 | 455, 1,465 |
| Low | 832 | 26, 1,637 | 772 | 266, 1,279 |
| Vendor food safety characteristics |  |  |  |  |
| Low | Ref | - | Ref | - |
| Moderate | 734 | -1,395, 2,863 | 508 | -137, 1,152 |
| High | -327 | -2,426, 1,771 | 248 | -363, 860 |
| Abbreviation: CI = Confidence Interval, TZS = Tanzanian Shillings. Notes: Models 1 and 2 were estimated using mixed-effect models with a random intercept at the village level, adjusting for covariates: household wealth, household produced any fruits and vegetables in the last 12 months, household dependency ratio; Women: age, education, occupation |  |  |  |  |

### 3.2. Qualitative

#### Women’s risk assessments

A brief summary of our field notes from observations and informal conversations that guided the interviews is provided in Supplemental T9. Here we present the results from the interviews with women.

Women sought to assess food safety risks by using sensory or visual cues from the produce while buying fruits and vegetables: checking for damage or deterioration, including signs of holes from insect or pest attacks; checking for spoilage, shrinkage, dryness, colour, and odour. A woman told us: *’When I select vegetables, I look for signs of freshness, such as a green colour and a firm texture. I also give them a sniff [smell] to check for any strange odours that might indicate spoilage or chemical contamination. Sometimes I will even chew a bit to get a sense of the taste and texture’* (26-30 years of age). Another woman mentioned ’..*unblemished fruit is preferable to bruised or damaged fruit, which may harbour insects’* (46-50 years of age). However, women also mentioned that excessive shine or unnatural colour made them wary of chemical contamination from pesticide residue. A vegetable vendor informed us that identifying pesticide contamination by appearance alone is difficult. She learned about this during a farmer’s training, along with the use of biopesticides (prepared from wild plants mixed with cow urine), which she believed kept the vegetables safer and longer-lasting than those treated with industrial pesticides.

Women in the communities complemented their visual or sensory assessments of risks by engaging with, observing, and developing a trustworthy relationship with the vendors. Participants reported asking vendors for a taste of fruit or vegetables, and sometimes vendors displayed and offered cut samples of fruits such as avocados, mangoes, and oranges. Women also used their observations of vendor food handling, hygiene and sanitation: ’*I look at whether the seller is cleaning fruits or has washed vegetables with water. If the environment he sells is cleaned or swept, and he has spread the vegetables on bags and not spread them down [on the earth floor]’* (46-50 years of age). Another woman who grew her own vegetables or occasionally bought them from neighbouring farms after a food-related illness in the family emphasised, ’*I have become more selective about where I buy [vegetables] from, opting for reputable vendors or sources known for their commitment to food safety*’ (36-40 years of age). Relatedly, when we asked women about their preferences for buying pre-cut fruit or vegetables, they reported concerns about vendors’ use of clean water for washing, personal hygiene, and cleanliness.

Women and vendors engaged in direct conversations about agrochemical contamination. At times, women denounced farmers for using chemicals, believing it was due to their greed to make ’fast money’. However, because they themselves lived in farming communities, they were also aware of the challenges farmers faced in controlling pests, dealing with harsh economic conditions, and adapting to weather changes. They also expressed understanding of the challenges vendors face in detecting contamination and maintaining proper hygiene. A woman told us: ’*I went to a farm just in my neighbourhood, and found a farmer who grows many vegetables to sell. I saw he grows his vegetables on the side [plot] for his family. I asked him why are you growing vegetables here on the side? He told me that these are for my family, and I do not spray them with pesticides. I asked him why are you spraying the rest? They are also consumed by humans, and you do that? He told me that they cannot grow without pesticides or fertiliser….*’ (46-50 years of age). Women vendors were aware of customers’ concerns about agrochemical contamination, so they showed empathy and practised positive reciprocity to address these concerns and retain loyalty. A woman vendor narrated, *’If a customer reports an issue like detecting pesticide odour in the vegetables they purchased, I would first listen to their concerns attentively and apologise for any inconvenience caused. Then, I would offer to refund their money or provide a replacement vegetable as compensation’* (36-40 years of age).

#### Learning mechanisms

Women reported learning about assessing food safety risks through their life experiences, particularly growing up in farming communities or families. A woman farmer told us: ’*I know the local old vegetables I grow have different smells compared to the sprayed ones. If you smell it, you will know it is different"* (46-50 years of age). Some women also mentioned being taught by their parents or in-laws, and having conversations with neighbours about how to detect and assess risks when buying fruit and vegetables. In addition, practising farming or selling gave the women access to training sessions, mostly on good agricultural practices or conducting business, offered by government or private institutions, horticulture organisations or village agricultural extension officers. A woman farmer, whose husband was also the chairman of a farmers’ group, told us: *’I am aware of the importance of following guidelines for proper pesticide usage. To stay informed, I have acquired knowledge from organisations like TAHA [*Tanzania Horticulture Association*] and also read the information provided on pesticide labels, where it is often indicated that vegetables should ideally be allowed to stay for 14 days after pesticide application. This practice ensures that the vegetables are safe for consumption and reduces any potential risks associated with pesticide residues’* (46-50 years of age). In sharp contrast, women in the Maasai community reported that the lack of agricultural activities in their village deprived them of the opportunity to learn about food safety risks through on-the-job training, and they rely on shared learning within the community or, sometimes, by observing women from other villages at the ward-level market. Hygiene practices, such as hand-washing, were also learned.

#### Risk consequences

Women across the three communities expressed concerns about the excessive use of agrochemicals, particularly synthetic pesticides in fruit and vegetable production. When we asked them about past food-related illnesses in the family, while a few of them recalled experiences of stomach aches, diarrhoea and vomiting, many said they did not, or that they had been ill after eating food, but not from fruit and vegetables. A couple of women farmers shared with us their awareness of the long-term health impacts of agrochemical contamination. One woman told us, ’*consuming contaminated fruit or vegetables treated with pesticides may not show immediate reactions, but the long-term effects can be detrimental, potentially leading to serious health issues like cancer. This underscores the importance of prioritising food safety and making informed choices about the products we consume*’ (41-45 years of age). Women also expressed helplessness while explaining the ubiquity of pesticides in fruit and vegetables and their effects.

#### Strategies to reduce risks

Despite fears of disease, several women believed they could control the risks by being vigilant when purchasing fruit and vegetables and by ensuring cleanliness during food preparation. To minimise the risk of purchasing unsafe fruit and vegetables, women reported modifying a range of purchasing behaviours, including rationing the quantity purchased, choosing certain varieties or quality believed to be safer, avoiding buying fruit and vegetables altogether, and altering their food sources. Women who were vegetable vendors recalled incidents in which their customers complained about the use of agrochemicals in their produce. After that, they either stopped buying from the market and started planting their own, or substituted it with a variety perceived as safer. Several women were willing to substitute commonly consumed green leafy vegetables, such as african nightshade, spinach, and amaranth, with pumpkin leaves, cowpea leaves, sweet potato leaves, and watercress, believing that these were typically grown without using agrochemicals and were also drought-resistant. One woman reported ’*I believe in buying from the fields [farms] that do not use pesticides and buy vegetables [variety] that do not need industrial fertilisers such as pumpkin leaves (’majani ya maboga’) and sweet potatoes leaves (’matembele’)’* (41-45 years of age). Another woman mentioned that she intentionally buys unripe fruit because she believes it is relatively safer than ripe fruit.

In villages one and two, where agro-climatic conditions supported agriculture, consuming fruits and vegetables from their farms was a common practice among women to ensure food safety and security, though it was seasonal. Women mentioned that their farms gave them control over both inputs and the variety to be planted. However, even within these villages, women relied on the market for vegetables during the dry season or when they did not have land to cultivate. One woman stated, ’*I decided to grow my own so that I can ensure safety. The decision to start growing [African] nightshades and amaranth came from a desire to take control of the quality and safety of the vegetables we eat. After learning about the potential risks associated with pesticides and other chemicals used in farming, I felt motivated to grow my own produce at home*’ (26-30 years old). Other women mentioned using ’natural fertilisers’ made from material such as neem leaves, cow urine, and ash.

Participants tackled food safety risks through hygiene, sanitation, handling, and storage practices at home. They mentioned washing with water (normal, running or warm), sorting, and clean storage as key household activities before cooking or eating. For fruits, they preferred not to store them overnight and to peel them before consumption. Cooking vegetables rather than eating them raw was a way women reported to kill bacteria. However, the women also noted that cooking needed to be done at an appropriate temperature to avoid destroying nutrients. A few of the women mentioned using plastic baskets for safe storage of fruits and vegetables away from insects, but most stored them on the floor in the household, covered with a cloth or sack. A woman also shared her preservation technique of sun-drying vegetables for extended usage: ’*I just take mixed types of vegetables, wash them, tie them in a plastic bag like those which are packed with sugar and boil them in a pan for a few minutes. Then I will dry them with direct sunlight. After it is dried, I will pack them and be ready for use, and I have used it for two weeks without any destruction to it*’ (36-40 years old). Finally, although none of the participants reported owning refrigerators, a few said that refrigeration kept food safe and extended its shelf life. Still, they were doubtful about how it influenced taste.

## 4. Discussion

In this analysis, we presented novel mixed-methods evidence on food safety from vendors in the food environment and from women in northern Tanzania. In our quantitative analysis, we found that women’s perceptions of food safety risks were negatively associated with household consumption and purchases of fruits and vegetables. However, village-level vendor food safety was not associated with these outcomes. Regarding food safety risk perceptions, women strongly agreed that buying from vendors who used hygienic and protective measures and choosing fruits and vegetables with a peel gave them control over food safety. In contrast, the excessive use of agrochemicals and preservation treatments in production and value chains heightened their concerns about food safety. Enumerator observations indicated that vendors had access to some infrastructure and implemented a few food safety practices, and that targeted support could further improve food safety. Our qualitative research revealed that women balanced their perceived control and concerns of food safety risks using a complex suite of food safety behaviours and practices embedded in their family and community lives. Women’s food safety risk assessments and related learnings were primarily derived from their social relations, in-person interactions, and trust in sellers. Additionally, women were assisted in minimising risk through their fluid and central role as producers, sellers, and at-home managers of fruit and vegetables.

This analysis contributes to the limited mixed-methods evidence on food safety as a driver of food choices for fresh foods in Africa (Isanovic et al., 2023; Karanja et al., 2022; Liguori et al., 2022; Stadlmayr et al., 2023). Our quantitative results showed that medium or lower food safety risk perceptions among women could influence consumption and expenditures by TZS 772-1,000 (person/month), which is ∼8-10% of households’ per-capita monthly consumption of fruit and vegetables. A rough comparison suggests that our estimated mean for per-capita monthly household fruit and vegetable consumption (TZS 11,454) was higher than the available estimate (TZS 9,375) for rural Tanzania households in 2017-18 (Rashid et al., 2024), which could be indicative of rising relative prices of fresh foods (Ignowski et al., 2023). However, the rural households in Tanzania are simultaneously buying and consuming more processed and ultra-processed foods and meals outside the home (Ignowski et al., 2023; Sauer et al., 2021). In this context, our results suggest that changing food safety perceptions could positively influence higher consumption and purchase of fresh fruits and vegetables, contributing to better diets. Other studies from urban or peri-urban East Africa have shown that food safety risk perceptions influenced the consumer behaviors of fresh foods (Bukachi et al., 2021; Downs et al., 2022; Kawemama et al., 2018; Lagerkvist et al., 2013; Pacho, 2020). Our results extend previous work by providing estimates for the perceived effect of perceptions on consumption and expenditure.

The food safety facilities and practices observed here were similar to those reported in other studies (Isanovic et al., 2023; Lee et al., 2022; Nordhagen, Lee, et al., 2022; Wallace et al., 2022). We found that vendors primarily lacked access to a washing station and access to a water source. Access to washing stations with clean potable water positively influences vendors’ personal hygiene (Wallace et al., 2022). It also ensures that fruit and vegetables are rinsed with uncontaminated water. However, merely installing washing stations without clean potable water or vendor awareness about pathogen transmission is likely to increase microbial contamination (Bello et al., 2011; Wallace et al., 2022). Relatedly, the surface and groundwater in northern Tanzania, which are used for washing and drinking purposes, have been previously found to be highly contaminated with inorganic pesticides and fertilisers and their application is increasing in the study regions (Lema et al., 2014; The United Republic of Tanzania, 2018, 2024). This also warrants attention to the quality of water used by approximately 50% of vendors who reported moistening of leafy vegetables. Providing safe water is also important from a consumer perspective, as one-half of our participants reported selecting items that were moistened by vendors. Lastly, dust, and dirt on and around the items were common, which can increase microbial contamination and escalate concern among consumers (Heilmann, 2016; Kumar et al., 2018; Lee et al., 2022; Rane, 2011). These findings suggest the need for programmes and interventions to improve facilities and to offer tailored training for vendors on effective waste management, food handling, and hygiene practices.

Our qualitative research revealed how social relations and trust enabled women to navigate food safety risks effectively. Trust is a well-established predictor of food safety risk perceptions, enabling consumers to exert control over their purchasing decisions (Ha et al., 2020; Machado Nardi et al., 2020). However, there is limited evidence on how trust is established and maintained in contexts with open, fresh food sources. We found that trust is not a unidimensional concept. That is, between the buyers and sellers, it embodied layers of actions and behaviours reproduced through talks, repeated exchanges, and reciprocity. Trust from a vendor’s perspective has been previously documented in studies from Africa, but mainly from a self-interest perspective of retaining customers (Lee et al., 2022; Rheinländer et al., 2008; Wenndt et al., 2025), or ’proprietor[s] … owing nothing to society …’ (Macpherson (1962: 3), as cited in (Busse, 2022)). However, the participants in this qualitative research often described learning to assess and trust food safety or quality from family, friends, neighbouring farmers or vendors. Our findings support the measurement of trust and social capital for a better understanding of consumer food safety risk assessment and mitigation (Lee et al., 2022; Nordhagen, Lambertini, et al., 2022). We also suggest developing culturally relevant, psychometrically validated measures of trust suitable for interactions between consumers and vendors in traditional food sources.

Despite the prevailing social attitudes towards food choices and safety, the qualitative responses also revealed a juxtaposition of moral values, conflictive and self-protective actions in food safety practices and behaviours. That is, for women, there is a fragile balance to maintain between harmony in social relations and ensuring safe consumption at home. Blake and colleagues noted that conflicting values persist in Tanzania and Kenya, primarily driven by the increasing availability of convenience or nontraditional foods, changing livelihoods, and strong intercultural and external influences (Blake et al., 2023). Our findings suggest that food safety knowledge interventions and information campaigns may be insufficient on their own; they need to address moral and social drivers. For example, vendors and consumers in a community could be brought together to find shared solutions that improve food safety.

Our findings indicate that food safety has implications for women’s diets, livelihoods, and their social and community relations. Other analyses using the FRESH evaluation data have shown that vegetables are a significant portion of women’s everyday diets (Azupogo et al., 2025), and indigenous varieties such as African eggplant, amaranth, spinach, and African nightshade, are purchased on a daily basis (Singh et al., 2025). However, our qualitative research suggested that due to food safety concerns, women’s preferences may be shifting away from commonly consumed and produced indigenous leafy vegetables, towards less widely available alternatives, such as sweet potato leaves or watercress (Kazungu, 2024; Singh et al., 2025). Previous research in another region of rural northern Tanzania has shown that purchase diversity is positively associated with dietary diversity of children (9-23 months) and their mothers (O’Malley et al., 2024). As an extension, higher food safety risk perceptions could worsen the current inadequate intake of fruit and vegetables among rural Tanzanian women (Amunga et al., 2024). Further investigation into farm and vendor safety practices of commonly consumed leafy vegetables is warranted for its implications on women’s nutrition and health. Longitudinal research can assist with monitoring of shifting perceptions and impacts on women’s diets.

Our findings demonstrated that the fluid positionalities of women, including their participation in activities across local fruit and vegetable food systems, constituted a key locus for understanding the social and material practices through which food safety knowledge is acquired and how women negotiate related risks. This suggests that efforts to improve food safety must be inclusive of their livelihoods, agency, time, and community relations, and situated knowledge (Barnett-Naghshineh, 2018; Lee et al., 2022; Waage et al., 2022). Previous studies have shown that the farm productivity of women in rural Tanzania is restricted by allocation and ownership of smaller plots in the family, lower levels of access to agricultural inputs, as well as earned incomes and limited support from men on their plots and in family responsibilities (Akram-Lodhi, 2024; van der Maden et al., 2021). Other studies have shown that lack of resources, support and control over income restrict women’s ability to mitigate food safety risks in their role as vendors and at home (Cook et al., 2024; Milovanova et al., 2024) Therefore, solutions to improve food safety or control hazards at the farm or vendor level should involve women and consider household situations and social norms (McMahon, 2013; McMahon & Glatt, 2018) Finally, interventions and programmes in socially embedded contexts like rural Tanzania can take guidance from the proposed ’three-legged stool’ theory of change model that combines individual-level awareness, training, and simple technology with community motivation, incentives, and an enabling macro environment (onboarding or support from regulatory authorities) (Grace, 2023). Lessons on facilitators and barriers should be drawn from the food safety planning and implementation of the animal-sourced foods in the country (Blackmore et al., 2022; MINISTRY OF LIVESTOCK AND FISHERIES, 2022; Waldman et al., 2020)

The Tanzanian government outlined a National Multisectoral Nutrition Action Plan for 2021-2026 to develop and strengthen food safety mechanisms and standards, thereby improving access to safe and nutritious fresh foods (United Republic of Tanzania, Prime Minister’s Office, 2021). However, the Plan’s outlook was primarily focused on formal markets in urban areas and traders, largely ignoring traditional sources, particularly those in rural or peri-urban areas (Kang’ethe et al., 2021). Simultaneously, the Arusha Sustainable Food System Platform aims to enhance the safe production and distribution of fresh foods at the city level through multi-stakeholder collaboration, including informal traders and consumers (Msuya, 2024). However, preliminary evidence suggests that the platform has faced difficulties in voicing or incorporating street vendors’ perspectives in a recent relocation decision (Steyaert et al., 2025). Our analysis demonstrates that food safety perceptions shaped by vendor practices in traditional markets and retail outlets is crucial for consumption of fruits and vegetables as well as for broader social relations so these sources should not be bypassed in improvement plans. Recent work is also recognising the role of traditional food markets or small traders in supporting urban diets and sustainability (UN Habitat, 2025). Relevant to our study context, we propose that efforts to improve food safety through interventions and governance should be conceptualised and implemented through a lens of inclusivity and equity, alongside protection (Cook et al., 2024; McMahon & Glatt, 2018). This can be achieved through a combination of interventions and programmes that target risk awareness and communication at the vendor and consumer level along with infrastructural support (Grace, 2023).

Regarding strengths, this research provides novel contextual evidence on food safety risk perceptions and their association with consumption and expenditure at the food group level. We also contribute to the methodology required for the social and cultural contextualisation of food safety by employing ethnographic methods. Our strategies to ensure that the qualitative research was rigorous, credible, and met the objectives included the use of local dialects of the tribes during data collection, familiarity of researchers with the food cultures in rural Tanzania in constructing the themes, regular peer reviewing of emerging themes individually and as a group, prolonged engagement in each community, and conducting follow-up interviews with the participants, a part of which involved sharing with them our preliminary themes. Ultimately, food safety encompasses multiple research disciplines, including public health, food science, toxicology, anthropology/ethnography, psychology, and economics. Therefore, multi-disciplinary mixed-methods approaches, similar to ours, are essential for future research and the design of people-centred, contextually appropriate interventions.

Our quantitative research was cross-sectional, and we cannot draw causal interpretations. However, mixed-methods approaches can serve as a relatively cost-effective and faster alternative to longitudinal studies for generating research evidence on food safety that informs interventions and policy (Wertheim-Heck & Raneri, 2019). While there is limited quantitative evidence on the subjective antecedents of food safety risk perceptions, such as perceived control and concerns, other known antecedents, such as trust and knowledge, could be included in future research to develop a comprehensive measure of food safety risk perceptions. This research combined individual, social, and food environment factors and did not focus on macro-level policy or governance mechanisms. Future studies could focus on the selection and incentives for local authorities in large markets, which have been shown to positively improve everyday issues such as garbage, dirt or dust (Resnick et al., 2025). Our qualitative research approached women’s perspectives on food safety primarily from their role as consumers of fresh foods. Still, we found that women often transgress the binaries of consumer, vendor, and producer in the agrifood system for fresh foods. Therefore, future qualitative work could offer extended perspectives on women and men as farmers or vendors, as well as their specific commodities. Finally, we recognise that food safety, as a driver of food choices for fruits and vegetables, has trade-offs and complementarities with other factors, such as freshness, accessibility, price, convenience, desirability, culture, and health. Hence, we have collected qualitative data on the broader drivers of food choices, which we aim to present in the future.

## Conclusion

This mixed-methods research highlighted that women’s perceptions of food safety were associated with household consumption and expenditure of fruit and vegetables in northern Tanzania. Women’s perspectives were influenced by their concerns about excessive agrochemical use in production, and food safety practices and facilities of the vendors. Women’s food safety risk assessments and related learnings were primarily derived from their social relations, community interactions, trust, and participation in food-related activities. Future research should build on the role of social and gender elements in the food environments to inform more effective interventions. Addressing food safety requires a combination of behavioural and structural solutions. The African Union and the Tanzanian government have a strong vision to produce and supply safe and nutritious food for all people, while ensuring sustainable livelihoods and economic growth (African Union, 2021; FAO, 2024). From a demand perspective, this research suggests that understanding food safety perceptions and practices within a broader community and food environment context is crucial for developing interventions to enhance choices for healthier foods and diets.

## Data Availability

All data produced in the present study are available upon reasonable request to the authors

## Ethics

Written informed consent was obtained from survey participants, including the head of the household, women, and vendors, before the start of each survey. Verbal consent was obtained from the owners for observations in the food environment. Ethical approval for the study was granted by the National Institute of Medical Research in Tanzania (NIMR/HQ/R.8a/Vol.IX/4357), the International Food Policy Research Institute (IFPRI) (00007490), Wageningen University and Research (2023-022), and the University of Edinburgh (HERC_2025_047). The qualitative research was approved by the University of Edinburgh (HERC_2023_183), the National Institute of Medical Research in Tanzania (NIMR/HQ/R.8a/Vol.IX/4357) and the International Food Policy Research Institute (IFPRI) (00007490). At the start of the qualitative interviews, we provided all the participants with an information sheet in Kiswahili that briefly outlined the study’s objectives, timing, confidentiality, and consent requirements.

## Author contributions

The authors’ responsibilities were as follows—Conceptualised the analyses: NS, QM, LB, ALB, DKO, MW, LMJ, NK Designed the FRESH study: LB, QM, NK, JK, DKO

Contributed to Quantitative data collection: NS, QM, LB, JLTC, EM, KJ, JK Contributed to Qualitative data collection: NS, MW, RZ, LM Quantitative data analysis: NS Qualitative data analysis: NS, RZ, LM Wrote the first version of the paper: NS Contributed to the first version of the paper: QM, ALB, MM, MW, LMJ, NK Commented on subsequent versions of the paper: all the co-authors Had primary responsibility for final content and all authors: NS, QM, MW, LMJ, NK Read and approved the final manuscript: all the co-authors

## Acknowledgements

We would like to thank Gayathri Ramani, Malick Dione, Rock Zagre, Wahid Quabili, Julia de Bryun, and the EDI Global team for their support in data management, cleaning, and analysis and Wiston Mwombeki for his support during fieldwork. We acknowledge Sonja Hess for her contribution to the study design.

## Corresponding author

Nishmeet Singh, PhD Scholar, University of Edinburgh

