## Supplemental Tables and Figures for "Women’s perceptions of food safety risks and vendor practices in Northern Tanzania: a mixed-methods analysis"

### Title

**Acknowledgements:**

We would like to thank Gayathri Ramani, Malick Dione, Rock Zagre, Wahid Quabili, Julia de Bryun, and the EDI Global team for their support in data management, cleaning, and analysis and Wiston Mwombeki for his support during fieldwork. We acknowledge Sonja Hess for her contribution to the study design.

**Corresponding author**

Nishmeet Singh, PhD Scholar, University of Edinburgh

**Keywords:** food safety, women's perceptions, vendor safety practices, fruits and vegetables, Africa, rural

### Supplemental

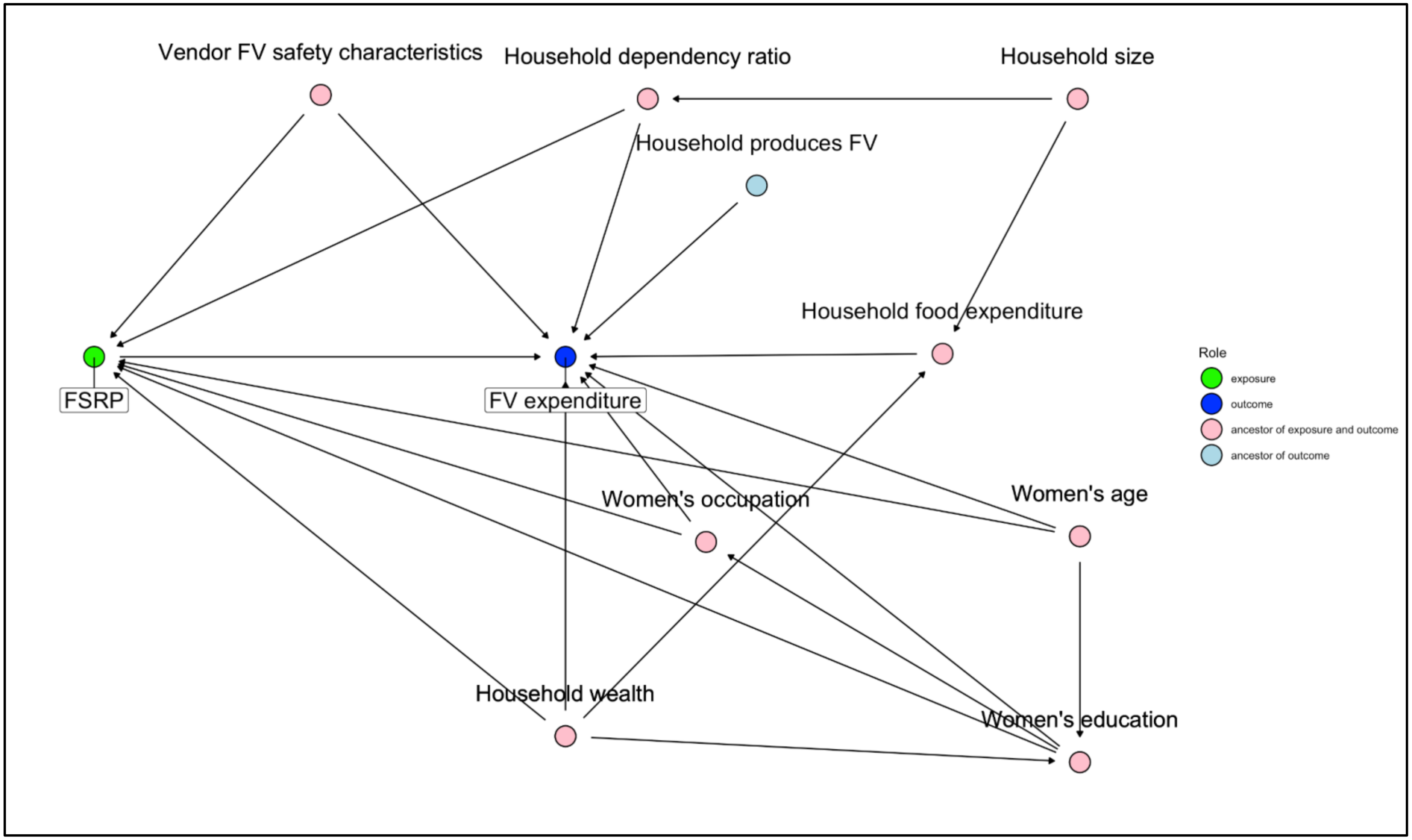

Sup Fig 1: Directed Acyclic Graph (DAG) map representing the causal pathways between food safety risk perception (FSRP) and fruit and vegetable (FV) consumption and expenditure, as well as potential confounders (Machado Nardi et al., 2020).

#### Supplemental Tables

Sup T1: Proportion of vendors with food safety facilities at the market (n=15) and retail outlet level (n=108) in October 2023

| | **Oct 2023** |
|---|---|
| **Markets' safety facilities score (out of 5),** median (Q1 - Q3) | 3.0 (2.0 - 4.0) |
| Dedicated waste collection area, % (n) | 93 (14) |
| Access to a nearby water source, % (n) | 60.0 (9) |
| Water station for handwashing or food washing, % (n) | 0 (15) |
| Closed sewage system % (n) | 66.7 (10) |
| Toilet within short walk, % (n) | 93.3 (14) |
| **Retail outlets' safety facilities score (out of 5),** median (Q1 - Q3) | 3.0 (3.0 - 4.0) |
| Dedicated waste collection area, % (n) | 82 (89) |
| Access to a nearby water source, % (n) | 75 (81) |
| Water station for handwashing or food washing, % (n) | 30 (32) |
| Closed sewage system, % (n) | 44 (47) |
| Toilet within short walk, % (n) | 98 (106) |

Notes: Based on enumerator observation data collected in the built food environments of rural Tanzania in October 2023

Sup T2: Proportion of vendors with fruit and vegetable safety practices at the markets (n=14) and retail outlets (n=107) across survey rounds

| | Average across rounds | Oct-23 | Nov-23 | Jan-24 |
|---|---|---|---|---|
| **Market safety practice score for fruit and veg (leafy and non-leafy) (Median (Q1 - Q3, out of 7)** | 4.6 (3.3 - 6.0) | NA | 4.6 (3.3 - 6.0) | NA |
| **Safety practice score for fruit (Median (Q1 - Q3, out of 7)** | 5.0 (4.0 - 7.0) | NA | 5.0 (4.0 - 7.0) | NA |
| **Fruit** | | | | |
| Garbage around the vendor's display | 53.8 (7) | NA | 53.8 (7) | NA |
| Dirty water around the vendor's display | 15.4 (2) | NA | 15.4 (2) | NA |
| Food items displayed on the ground | 46.2 (6) | NA | 46.2 (6) | NA |
| Dust or dirt on the food items | 38.5 (5) | NA | 38.5 (5) | NA |
| Dust or dirt around the display of food items | 23.1 (3) | NA | 23.1 (3) | NA |
| Food items exposed to direct sunlight or heat | 30.8 (4) | NA | 30.8 (4) | NA |
| Food items exposed to rain | 30.8 (4) | NA | 30.8 (4) | NA |
| Food items moistened using water in a spray bottle or other vessel | 0.0 (1) | NA | 0.0 (1) | NA |
| Food items moistened by rinsing them in an open bowl or jug of water | 15.4 (2) | NA | 15.4 (2) | NA |
| Vendor ever sell food items that are already cut or peeled | 30.8 (4) | NA | 30.8 (4) | NA |

| | | | | |
|---|---|---|---|---|
| **Safety practice score for Non-leafy vegetables (Median (Q1 - Q3, out of 8)** | 5.0 (3.0 - 6.0) | NA | 5.0 (3.0 - 6.0) | NA |
| **Non-leafy vegetables** | | | | |
| Garbage around the vendor's display | 42.9 (6) | NA | 42.9 (6) | NA |
| Dirty water around the vendor's display | 7.1 (1) | NA | 7.1 (1) | NA |
| Food items displayed on the ground | 57.1 (8) | NA | 57.1 (8) | NA |
| Dust or dirt on the food items | 28.6 (4) | NA | 28.6 (4) | NA |
| Dust or dirt around food item's display | 50.0 (7) | NA | 50.0 (7) | NA |
| Food items exposed to direct sunlight or heat | 35.7 (5) | NA | 35.7 (5) | NA |
| Items exposed to rain | 28.6 (4) | NA | 28.6 (4) | NA |
| Food items moistened using water in a spray bottle or other vessel | | NA | | NA |
| Food items moistened by rinsing them in an open bowl or jug of water | 100.0 (14) | NA | 100.0 (14) | NA |
| Vendor ever sell food items that are already cut or peeled | 21.4 (3) | NA | 21.4 (3) | NA |
| **Safety practice score for Leafy vegetables (Median (Q1 - Q3, out of 8)** | 4.0 (3.0 - 6.0) | NA | 4.0 (3.0 - 6.0) | NA |
| **Leafy vegetables** | | | | |
| Garbage around the vendor's display | 61.5 (8) | NA | 61.5 (8) | NA |
| Dirty water around the vendor's display | 38.5 (5) | NA | 38.5 (5) | NA |
| Food items displayed on the ground | 38.5 (5) | NA | 38.5 (5) | NA |

| | | | | |
|---|---|---|---|---|
| Dust or dirt on the food items | 38.5 (5) | NA | 38.5 (5) | NA |
| Dust or dirt around the display of food items | 15.4 (2) | NA | 15.4 (2) | NA |
| Food items exposed to direct sunlight or heat | 23.1 (3) | NA | 23.1 (3) | NA |
| Food items exposed to rain | 23.1 (3) | NA | 23.1 (3) | NA |
| Food items moistened using water in a spray bottle or other vessel | 23.1 (3) | NA | 23.1 (3) | NA |
| Food items moistened by rinsing them in an open bowl or jug of water | 46.2 (6) | NA | 46.2 (6) | NA |
| Vendor ever sell food items that are already cut or peeled | 46.2 (6) | NA | 46.2 (6) | NA |
| **Retail outlet safety practice score (Median (Q1 - Q3, out of 7)** | 6.0 (5.0 - 7.0) | 7.0 (5.0 - 7.0) | 7.0 (6.0 - 7.0) | 6.0 (4.0 - 7.0) |
| Garbage around the outlet, % (n) | 20.8 (47) | 21.8 (17) | 15.4 (12) | 25.7 (18) |
| Dirty water around the outlet, % (n) | 6.2 (14) | 1.3 (1) | 10.3 (8) | 7.1 (5) |
| Fruits and vegetables displayed on the ground, % (n) | 10.4 (23) | 6.6 (5) | 10.4 (8) | 14.5 (10) |
| Dust or dirt on fruits and vegetables, % (n) | 25.7 (57) | 28.9 (22) | 19.5 (15) | 29.0 (20) |
| Dust or dirt around fruits and vegetables, % (n) | 30.6 (68) | 27.6 (21) | 20.8 (16) | 44.9 (31) |
| Fruits and vegetables exposed to direct sunlight or heat, % (n) | 4.5 (10) | 3.9 (3) | 3.9 (3) | 5.8 (4) |
| Fruits and vegetables exposed to rain, % (n) | 3.6 (8) | 2.6 (2) | 2.6 (2) | 5.8 (4) |
| Fruits and vegetables moistened using water in a spray bottle, % (n) | 14.6 (32) | 10.8 (8) | 17.1 (13) | 15.9 (11) |

| | | | | |
|---|---|---|---|---|
| Fruits and vegetables moistened by rinsing them in an open vessel, % (n) | 38.0 (84) | 35.5 (27) | 35.5 (27) | 43.5 (30) |
| Outlet ever sell fruits and vegetables that are already cut or peeled, % (n) | 32.0 (71) | 23.4 (18) | 34.2 (26) | 39.1 (27) |

Notes: Based on enumerator observation data collected in the built food environments of rural Tanzania. The retail outlets were observed for three rounds in October 2023, November 2023 and January 2024, while the market vendors were observed in November 2023.

Sup T3: Demographic characteristics of the women (respondent) and the household (n=2,577)

| | **Mean (SD); %(n)** |
|---|---|
| Respondent characteristics | |
| Age (y) | 38.3 (6.2) |
| Highest level of education | |
| *Primary incomplete* | 6.4% (164) |
| *Primary complete* | 66.5% (1,714) |
| *Secondary incomplete* | 3.4% (87) |
| *Secondary complete* | 8.8% (227) |
| *Higher* | 2.9% (74) |
| *Never went to school* | 12.1% (311) |
| Primary activity (majority of time spent), last 12 m | |
| *Self-employed in agricultural activity (farming/livestock)* | 59.3% (1,529) |
| *Employed in a non-food-related activity* | 24.7% (636) |
| *Employed in food system activity* | 7.5% (193) |
| *Other (Unemployed/not seeking work/student)* | 4.2% (109) |
| *Household work, including childcare* | 4.3% (110) |
| Marital Status | |

| | |
|---|---|
| *Never married* | 3.5% (91) |
| *Married or living together* | 85.5% (2,203) |
| *Widowed/divorced/separated* | 11.0% (283) |
| HH produced any fruit and vegetable, last 12 m | 36.4% (937) |
| Wealth index | |
| *Lowest* | 20.0% (516) |
| *Lower* | 19.9% (514) |
| *Middle* | 20.0% (516) |
| *Higher* | 20.0% (515) |
| *Highest* | 20.0% (516) |
| Household characteristics | |
| Household size | 5.8 (1.7) |
| Household dependency ratio | 1.2 (0.8) |

Sup T4: Description of women's fruit and vegetable safety risk perceptions (Oct 2023- Jan 2024, n=2593)

| **Food safety risk perception score out of 110, median (Q1 - Q3 )** | 58 (52 - 65) | | | | |
|---|---|---|---|---|---|
| | **High risk perception** | → | | | **Low risk perception** |
| **Risk perception statements** | **Completely Disagree (1)** | **Disagree (2)** | **Neutral (3)** | **Agree (4)** | **Completely Agree (5)** |
| **Fruits** | | | | | |
| 1. I buy those fruits that my vendor advises me to buy | 18% | 39% | 9.7% | 25% | 7.7% |
| 2. I only buy fruits from vendors that use hygienic and protective measures (i.e. no dust, dirt, rotting garbage, dirty water, direct sun) | 1.3% | 3.0% | 4.3% | 52% | 39% |
| 3. I only buy fruits when I know where they are produced | 24% | 43% | 14% | 17% | 2.2% |
| 4. I only buy when the fruits are moistened using safe potable water contained in a spray bottle | 16% | 17% | 16% | 37% | 14% |
| 5. I only select fruits that carry food safety certification or labels | 28% | 40% | 14% | 17% | 1.1% |
| 6. I only select fruits that have a peel or are uncut | 3.9% | 3.0% | 1.4% | 43% | 49% |
| **Vegetables** | | | | | |
| 1. I buy those vegetables that my vendor advises me to buy | 17% | 38% | 9.9% | 26% | 9.6% |

| | | | | | |
|---|---|---|---|---|---|
| 2. I only buy vegetables from vendors that use hygienic and protective measures (i.e. no dust, dirt, rotting garbage, dirty water, direct sun) | 0.5% | 2.2% | 5.0% | 51% | 41% |
| 3. I only buy vegetables when I know where they are produced | 17% | 36% | 13% | 26% | 9.1% |
| 4. I only buy when the vegetables are moistened using safe potable water contained in a spray bottle | 15% | 17% | 15% | 39% | 14% |
| 5. I only select vegetables that carry food safety certification or labels | 30% | 44% | 13% | 13% | 0.8% |
| 6. I only select vegetables that have a peel or are uncut | 5.0% | 3.0% | 1.2% | 42% | 49% |
| **Perceived concern statements** | **Low risk perception** | → | | | **High risk perception** |
| How concerned are you about the following: | **Totally not concerned (5)** | **Not so concerned (4)** | **Neutral (3)** | **Rather concerned (2)** | **Totally concerned (1)** |
| **Fruits** | | | | | |
| Fertilisers and growth enhancers | 12% | 27% | 12% | 15% | 34% |
| Pesticides | 10% | 23% | 10% | 13% | 43% |
| Preservation treatment | 3.7% | 16% | 9.3% | 18% | 53% |
| Unhygienic handling practices (store/vendor) | 2.9% | 19% | 10% | 15% | 54% |
| Use of /growing with contaminated water and soil | 6.5% | 25% | 15% | 14% | 39% |

| **Vegetables** | | | | | |
|---|---|---|---|---|---|
| Fertilisers and growth enhancers | 7.8% | 22% | 7.3% | 17% | 46% |
| Pesticides | 5.0% | 15% | 4.9% | 14% | 61% |
| Preservation treatment | 2.4% | 13% | 6.1% | 17% | 61% |
| Unhygienic handling practices (store/vendor) | 1.7% | 16% | 7.2% | 16% | 59% |
| Use of /growing with contaminated water and soil | 3.4% | 22% | 12% | 16% | 46% |

Sup T5: Summary of monthly per capita fruits and vegetables consumption expenditures and purchases in Tanzanian Shillings (TZS)

| Characteristic | n = 2,577 |
|---|---|
| Fruits and Vegetables consumption expenditures (TZS/person/month) | |
| Mean (SD) | 11,454 (9,196) |
| Median (Q1, Q3) | 9,283 (5,138, 14,877) |
| Min, Max | 0, 75,915 |
| Fruits and Vegetables purchases (TZS/person/month) | |
| Mean (SD) | 7,298 (5,746) |
| Median (Q1, Q3) | 5,866 (3,205, 9,622) |
| Min, Max | 0, 39,107 |

Sup T6: Table of correlation analysis between raw scores of women’s food safety risk perceptions and village-level vendor safety characteristics (n=2,577)

Kendall's rank correlation tau estimate: -0.011
z = 0.84214, p-value = 0.3997

Sup T7: Association between women's food safety risk perceptions, vendor food safety, and and households’ combined fruit and vegetable consumption (n=2577)

Dependent variable: Combined fruit and vegetable per capita monthly household consumption (TZS)

| **Independent variable** | **Beta** | **95% CI** | **p-value** |
|---|---|---|---|
| Women’s risk perceptions | | | |
| High | Ref | — | |
| Medium | 1,000 | 194, 1,805 | 0.015 |
| Low | 832 | 26, 1,637 | 0.043 |
| Vendor food safety characteristics | | | |
| Low | Ref | — | |
| Moderate | 734 | -1,395, 2,863 | 0.5 |
| High | -327 | -2,426, 1,771 | 0.8 |
| Household wealth quintile | | | |

| | | | |
|---|---|---|---|
| Lowest | Ref | — | |
| Lower | 127 | -967, 1,221 | 0.8 |
| Middle | 1,342 | 197, 2,487 | 0.022 |
| Higher | 2,425 | 1,223, 3,627 | <0.001 |
| Highest | 5,246 | 3,976, 6,515 | <0.001 |
| Household produced any fruit and vegetables, last 12 m | 1,687 | 911, 2,463 | <0.001 |
| Household member's age | 12 | -45, 69 | 0.7 |
| What is the highest level of education? | | | |
| Primary incomplete | Ref | — | |
| Primary complete | 518 | -906, 1,943 | 0.5 |
| Secondary incomplete | 1,369 | -898, 3,636 | 0.2 |
| Secondary complete | 1,947 | 137, 3,757 | 0.035 |
| Higher | 1,786 | -683, 4,256 | 0.2 |
| Never went to school | -495 | -2,100, 1,109 | 0.5 |
| What was the primary labour activity (majority of time spent) in the last 12 mon | | | |
| Self-employed in agriculture activity (farming/livestock) | Ref | — | |
| Employed in non-food related activity | 737 | -184, 1,658 | 0.12 |
| Employed in food system activity | 558 | -759, 1,876 | 0.4 |

| | | | |
|---|---|---|---|
| Other (Unemployed/not seeking work/student) | -408 | -2,126, 1,309 | 0.6 |
| Household work including children | -2,314 | -3,993, -634 | 0.007 |
| Household dependency ratio | -132 | -591, 327 | 0.6 |
| village.SD (Intercept) | 2,286 | | |

Abbreviation: CI = Confidence Interval

Sup T8: Association between combined fruit and vegetable expenditures and vendor food safety and women's food safety risk perceptions

Dependent variable: Fruit and Vegetable per capita monthly household expenditure (TZS)

| **Independent variable** | **Beta** | **95% CI** | **p-value** |
|---|---|---|---|
| Women’s risk perceptions | | | |
| High | Ref | — | |
| Medium | 960 | 455, 1,465 | <0.001 |
| Low | 772 | 266, 1,279 | 0.003 |
| Vendor food safety characteristics | | | |
| Low | Ref | — | |
| Moderate | 508 | -137, 1,152 | 0.12 |
| High | 248 | -363, 860 | 0.4 |

| | | | |
|---|---|---|---|
| Household wealth quintile | | | |
| Lowest | Ref | — | |
| Lower | 610 | -66, 1,286 | 0.077 |
| Middle | 1,103 | 396, 1,809 | 0.002 |
| Higher | 2,578 | 1,845, 3,311 | <0.001 |
| Highest | 4,499 | 3,731, 5,267 | <0.001 |
| hh_prodfv | -62 | -523, 399 | 0.8 |
| Household member's age | -28 | -63, 8.0 | 0.13 |
| What is the highest level of education? | | | |
| Primary incomplete | Ref | — | |
| Primary complete | 547 | -339, 1,434 | 0.2 |
| Secondary incomplete | 95 | -1,329, 1,519 | 0.9 |
| Secondary complete | 1,643 | 508, 2,777 | 0.005 |
| Higher | 1,208 | -346, 2,761 | 0.13 |
| Never went to school | -70 | -1,078, 939 | 0.9 |
| What was the primary labour activity (majority of time spent) in the last 12 mon | | | |

| | | | |
|---|---|---|---|
| Self-employed in agriculture activity (farming/livestock) | Ref | — | |
| Employed in non-food related activity | 1,177 | 617, 1,738 | <0.001 |
| Employed in food system activity | 586 | -231, 1,402 | 0.2 |
| Other (Unemployed/not seeking work/student) | -395 | -1,460, 671 | 0.5 |
| Household work including children | -1,130 | -2,178, -83 | 0.034 |
| Household dependency ratio | -330 | -619, -41 | 0.025 |
| village.SD (Intercept) | 399 | | |

Abbreviation: CI = Confidence Interval

Sup T9: Summary of field notes

From our observations and informal interviews, conducted prior to the formal interviews, we noted similarities and differences in the everyday lives of women, food cultures, and the food sources, which helped us contextualise food safety. Households primarily utilised ward-level weekly markets to buy fruits and vegetables. Other independent sources, such as kiosks, retail shops, mobile vendors, or direct purchases from farms were used for vegetables. We observed that the physical infrastructure in the markets consisted mainly of vast, open areas under direct sunlight, without an overarching roof, and fruits and vegetables were displayed by vendors under private umbrellas on top of jute sacks on a natural earth or earth floor. However, some markets also featured spaces with several vendors under a permanent tin roof, a concrete floor, as well as vendors with individual wooden kiosks or tables.

In the three sampled villages, women were at the core of decisions regarding fruit and vegetables, including growing, selling, and preparing, but differences were observed between them, which could influence food safety perspectives and food choices For example, women in the first village were growing fruit and vegetables, for home consumption and selling due to suitable agro-climatic conditions, fertile soils, and proximity to the town market. In contrast, women in the third village were becoming relatively disengaged from direct food production. Instead, they preferred to sell fruit and vegetables, managed retail stores (Duka) stocked with foods like peanut butter, pasta, and tomato ketchup, worked in restaurants or hotels, or were employed as helpers in houses or beauty salons. A possible reason mentioned was the demand from the nearby foreign residential area and the rise of tourism. The third village was a pastoralist ‘Maasai’ (tribe) community and farthest from the town. It had very limited agriculture activities due to the traditional occupation of livestock raising and unsuitable agro-climatic conditions (water scarcity and rocky, dry, and semi-arid land).
